# Blood donor exposome and impact of common drugs on red blood cell metabolism

**DOI:** 10.1101/2020.08.17.20176891

**Authors:** Travis Nemkov, Davide Stefanoni, Aarash Bordbar, Aaron Issaian, Bernhard O. Palsson, Larry J Dumont, Ariel Hay, Anren Song, Yang Xia, Jasmina S. Redzic, Elan Z. Eisenmesser, James C Zimring, Steve Kleinman, Kirk C. Hansen, Michael P. Busch, Angelo D’Alessandro, for the Recipient Epidemiology and Donor Evaluation Study-III (REDS-III) RBC-Omics Study

## Abstract

Computational models based on recent maps of the red blood cell proteome suggest that mature erythrocytes may harbor targets for common drugs. This prediction is relevant to red blood cell storage in the blood bank, in which the impact of small molecule drugs or other xenometabolites deriving from dietary, iatrogenic or environmental exposures (“exposome”) may alter erythrocyte energy and redox metabolism and, in so doing, affect red cell storage quality and post-transfusion efficacy. To test this prediction, here we provide a comprehensive characterization of the blood donor exposome, including the detection of common prescription and off-the-counter drugs in 250 units donated by healthy volunteers from the REDS-III RBC Omics study. Based on high-throughput drug screenings of 1,366 FDA-approved drugs, we report a significant impact of ∼65% of the tested drugs on erythrocyte metabolism. Machine learning models built using metabolites as predictors were able to accurately predict drugs for several drug classes/targets (bisphosphonates, anticholinergics, calcium channel blockers, adrenergics, proton-pump inhibitors, antimetabolites, selective serotonin reuptake inhibitors, and mTOR) suggesting that these drugs have a direct, conserved, and significant impact on erythrocyte metabolism. We then focused on ranitidine – a common antiacid – as a representative drug with the potential to improve human erythrocyte storage quality and post-transfusion performances in mice. By combining tracing experiments with 1,2,3-^13^C_3_-glucose, proteome integral solubility alteration assays, genetic ablation of S1P synthesis capacity, *in silico* docking and 1D NMR, we show that ranitidine triggers metabolic mechanisms involving sphingosine 1-phosphate (S1P)-dependent modulation of erythrocyte glycolysis and/or direct binding to hemoglobin.

**Graphical Abstract:** RBC exposome from the REDS III study revealed that blood from a subset of donors contains traces of the most common drugs in the United States. RBCs can uptake these drugs, in some cases can metabolize them to their bioactive metabolites and in others the drug can directly impact RBC metabolism during storage.

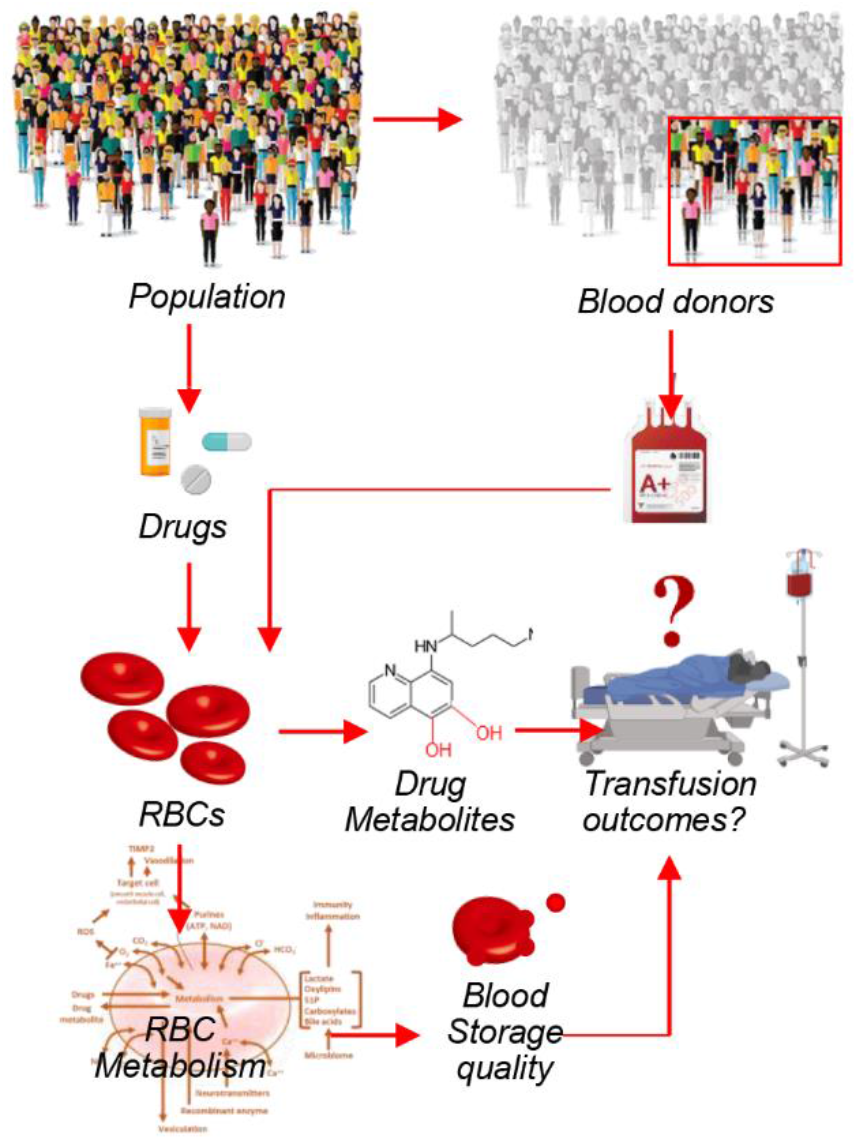

**Key points:**

1. Blood donor exposomes include metabolites of environmental exposure, traces of common prescription or off-the-counter drugs;
2. 65% of 1366 FDA- approved drug significantly affect RBC metabolism. Ranitidine significantly impacts glycolysis and S1P metabolism.

## Introduction

Despite being the most abundant cell type in the human body,^1^ red blood cells (RBCs) are usually regarded as a bystander in systems physiology. RBCs play a critical role in the transport and delivery of oxygen to tissues, a task that they have evolved to fulfill by maximizing the oxygen carrying capacity: having lost nuclei and organelles, the mature RBC carries ∼250–270 million molecules of hemoglobin per cell.^2^ As such, most textbooks refer to the mature erythrocyte as a mere circulating bag of hemoglobin that serves no other purpose. Two decades of advances in the field of proteomics have revealed that – while ∼90% of the dry weight of a mature erythrocyte is indeed comprised of hemoglobin – the residual 10% harbors a rich and diverse protein machinery of approximately 3,000 gene products.^3^ Several studies have shown that the mature RBC is capable of responding to environmental stimuli (e.g., hypoxia and oxidant stress) through several mechanisms that involve post-translational modifications (especially phosphorylation^4^ and methylation^5^) and metabolic reprogramming. These cascades are usually triggered by direct stimulation of receptors on the cell surface (e.g., adenosine receptor in response to high-altitude hypoxia^6^) or hemoglobin-oxygen saturation, which affects the compartmentalization and activity of several metabolic (e.g. glyceraldehyde 3-phosphate dehydrogenase^7,8^) and antioxidant enzymes (e.g. peroxiredoxin 2^9^). These mechanisms of oxygen-dependent metabolic modulation also promote a feedback in energy and redox metabolism, closing a loop that ultimately affects the capacity of RBCs to capture, carry and off-load oxygen. Some of these receptors include purinergic, muscarinic, dopaminergic or other neurotransmitter-stimulated receptors,^10^ making the mature RBC theoretically sensitive to small molecules that act as agonist or antagonist of those signaling pathways in a way that could ultimately affect RBC metabolism and, in so doing, hemoglobin capacity to bind and off-load oxygen.

The unanticipated complexity of the RBC proteome suggests that the mature erythrocyte could not only respond upon stimulation of specific drug targets – *in silico* predictions suggest that ∼230 targets to common drugs may exist in the mature RBC^11^ – but could also theoretically participate in drug metabolism. Some of the proteins compiled in the latest proteomics studies,^3,12^ if functional, could play a role in cellular – and maybe systems – metabolism. For example, recent studies have shown that RBCs are endowed with several transporters, as well as diaphorases and enzymes with cytochrome b5 reductase activity^13^, which allows them to metabolize antimalarial drugs such as chloroquine^14^ or antiretroviral drugs (e.g., tenofovir^15^).

In recent years, the introduction of omics technologies to the field of transfusion medicine has expanded our understanding of the mechanisms underlying the progression and severity of the storage lesion^16^. While blood storage is a logistic necessity to make over 100 million units available for transfusion every year worldwide, refrigerated storage in the blood bank is accompanied by the progressive accumulation of storage lesions^17^. Oxidant stress to metabolic enzymes, structural and functional proteins^5,8,18^ ultimately results in RBCs that are more susceptible to lysis in the bag or untimely removal from the bloodstream in the recipient. The rate at which the storage lesions occur and the extent to which these phenomena progress by the end of the shelf-life of a blood unit (42 days in the United States) vary depending on processing strategies, storage bags and solutions and donor biology.^19–21^ These factors have emerged clearly from the Recipient Epidemiology and Donor Evaluation Study (REDS-III) RBC-Omics Study, in which 13,403 healthy whole blood donor volunteers consented to participate on a large scale assessment of donor-dependent factors on RBC storability and hemolytic propensity at the end of storage. Additional factors such as sex, ethnicity, age,^20^ donation frequency or genetic polymorphisms are associated with a decreased capacity of the mature RBC to cope with osmotic and oxidant stress (e.g., glucose 6-phosphate dehydrogenase deficiency), and ultimately affect the “metabolic age” and post-transfusion efficacy of the unit.^22,23^

It is a logical consequence of the studies described above that the antioxidant capacity of a mature RBC is affected by other factors beyond donor biology that could counteract or aggravate oxidant stress. One such factor is represented by donor habits, such as smoking, which in smaller cohorts has been shown to increase markers of oxidant stress in the stored unit (e.g., carboxyhemoglobin or altered glutathione homeostasis^24^) and ultimately result in lower hemoglobin increments in recipients of units from smoking donors^25^. However, it is unclear whether and to what extent other factors derived from environmental exposures – referred to as the “exposome”^26^ – would impact the metabolic age of the stored RBC and, as a result, transfusion efficacy in the recipient.

## Methods

Extensive details are provided in the **Supplementary File – Material and Methods extended**.

### REDS-III RBC-Omics study participants and samples

Donor selection and recruitment for the RBC-Omics study under approved protocols (BioLINCC Study: HLB02071919a) were previously detailed.^27–29^ Samples from 13,403 RBC units were stored for ∼39–42 days, prior to evaluation for osmotic^30,31^ and oxidative hemolysis^30^ (12,799 and 10,476, respectively). A subset (n = 250) of extreme hemolyzers in this cohort (5^th^ and 95^th^ percentile) donated a second unit, which was sampled at storage day 10, 23 and 42. A subset of these samples (599 total samples) were made available for metabolomics via ultra-high-pressure liquid chromatography coupled to high resolution mass spectrometry (UHPLC-MS – Vanquish-QExactive, Thermo Fisher, San Jose, CA, USA).^30,31^

### High-throughput drug screening

Three leukocyte-filtered units from healthy donor volunteers were collected in CP2D-AS-3 and incubated with a screening library of 1,366 Food and Drug Administration (FDA)-approved drugs at 10 μM concentration for 24h at 37 under sterile conditions in 96 well plates. Automated extractions and high-throughput metabolomics analyses are detailed in previous technical papers^32^ and in the ***Supplementary File***.^33,34^

### Incubation with ranitidine of Human and Mouse RBCs

RBCs from blood donors or mice – either WT or Sphk1 KO, as described^35^ were incubated in CPD-AS-3 or CPDA1, respectively, with increasing doses (10, 25, 50, 100, 200 uM) of ranitidine (product no: 1598405 – Millipore Sigma), in presence of 1,2,3-^13^C_3_-glucose (product no: 720127 – SIGMA Aldrich, St Louis, MO, USA) to determine fluxes through glycolysis based on the isotopologue M+3 of lactate.^8^

### Post-transfusion recovery of mouse RBCs stored in presence of ranitidine

Mouse RBCs were obtained by intracardiac puncture from wild-typeFVB/J and C57BL6/J mice (The Jackson Laboratory).

All mice were housed in the University of Virginia vivarium, and all procedures were performed under an Institutional Animal Care and Use Committee–approved protocol. RBCs were stored in CPDA1 additive,^36^ either untreated or supplemented with 50, 100, 200 uM ranitidine for up to 8 days at 4ºC in at least three independent experiments (n = 3 per group), prior to determination of post-transfusion recoveries^36^ (***Supplementary File)***.

### Proteome Integral Solubility Alteration (PISA) assay

PISA experiments were performed as described extensively in methodological papers,^37,38^ by determining temperature-dependency of protein solubility after incubating human RBCs or A549 epithelial cells (n = 5) in presence or absence of ranitidine 100 uM in the interval of 43–57ºC. Protein fractions underwent TMT-labeling, high-pH reversed phase fractionation and nanoUHPLC-MS/MS proteomics (Orbitrap Fusion Lumos, Thermo Fisher, Bremen, Germany).

### 1D Nuclear Magnetic Resonance assay of ranitidine and hemoglobin interaction

1D NMR 1H spectrum was collected at 25ºC for 200 uM ranitidine alone and in the presence of 20 or 200 uM human hemoglobin tetramers (product no: H7379–1G – SIGMA Aldrich, St Louis, MO, USA). A total of 48 scans were collected on a Varian 900 using the BioPack water sequence implemented with wet water suppression.

### Statistical Analyses

Graphs and statistical analyses (either t-test or repeated measures ANOVA) were prepared with GraphPad Prism 5.0 (GraphPad Software, Inc, La Jolla, CA) and MetaboAnalyst 4.0. Drug library screening data were processed via t-distributed stochastic neighbor embedding (TsNE) to highlight the extent of metabolic impact of drug exposure on RBC metabolism and biclustering analysis to identify specific metabolic targets for subset of drugs.

## Results

### Identification of xenometabolites in the blood of healthy donor volunteers

Blood units from 250 healthy donor volunteers were sampled at storage days 10, 23 and 42 for metabolomics analyses of the RBC exposome, i.e. the compendium of small molecule metabolites derived from diet, the microbiome or other xenometabolites, such as pollutants or drugs^26^ (**Figure 1.A –** data in **Supplementary Table 1**). To ensure comprehensiveness of the analysis, a custom database was imported from the *Blood Exposome* website (**Supplementary Table 1**).^39^ Unsupervised principal component analyses (PCA) clustered the subjects on the basis of storage duration (red, green, blue for days 10, 23 and 42, respectively – across PC1) and additive solution (across PC2 – **Figure 1.B**). Hierarchical clustering analysis of the exposome data (**Figure 1.C**) highlighted groups of donors with unique metabolic status and various habits (**Figure 1.D-H**). Dot plots in **Figure 1.I-M** provide an overview of the levels of the most abundant exposome metabolites of microbial origin, derived from donor habits (e.g., drinking alcohol or caffeinated beverages; nicotine exposure), plasticizers (e.g., from blood bags) and dietary metabolites. Of note, when searching against the list of the 200 most common prescription or off-the-counter drugs in the United States,^40^ we could detect traces of most compounds in at least one donor – with the most frequently detected compounds listed in **Figure 1.N (**full report in **Supplementary Table 1**). For example, traces of drugs detected in RBC units included angiotensin II receptor inhibitors, NSAID and anti-inflammatory drugs, antihypertensive, pain medications, antidepressants and sleep aid (**Figure 2**), drugs targeting specific metabolic enzymes, such as the rate-limiting enzyme in cholesterol synthesis – hydroxymethyl-glutaryl CoA reductase inhibitors – in the case of statins, or systems metabolism at large (e.g., biguanides like metformin – **Supplementary Figure 1**). Three units from subjects > 70 years of age tested positive for sildenafil – an inhibitor of the enzyme phoshodiesterase E5 that regulates cGMP metabolism – and its catabolite desmethyl-sildenafil (**Supplementary Figure 2**). Sildenafil was associated with increased level of oxidative stress-related metabolites and increased susceptibility to oxidant stress-induced hemolysis, but lower osmotic fragility, as gleaned by unsupervised PCA, hierarchical clustering and pathway analyses(**Supplementary Figure 2.E-G**).

**Figure 1.**
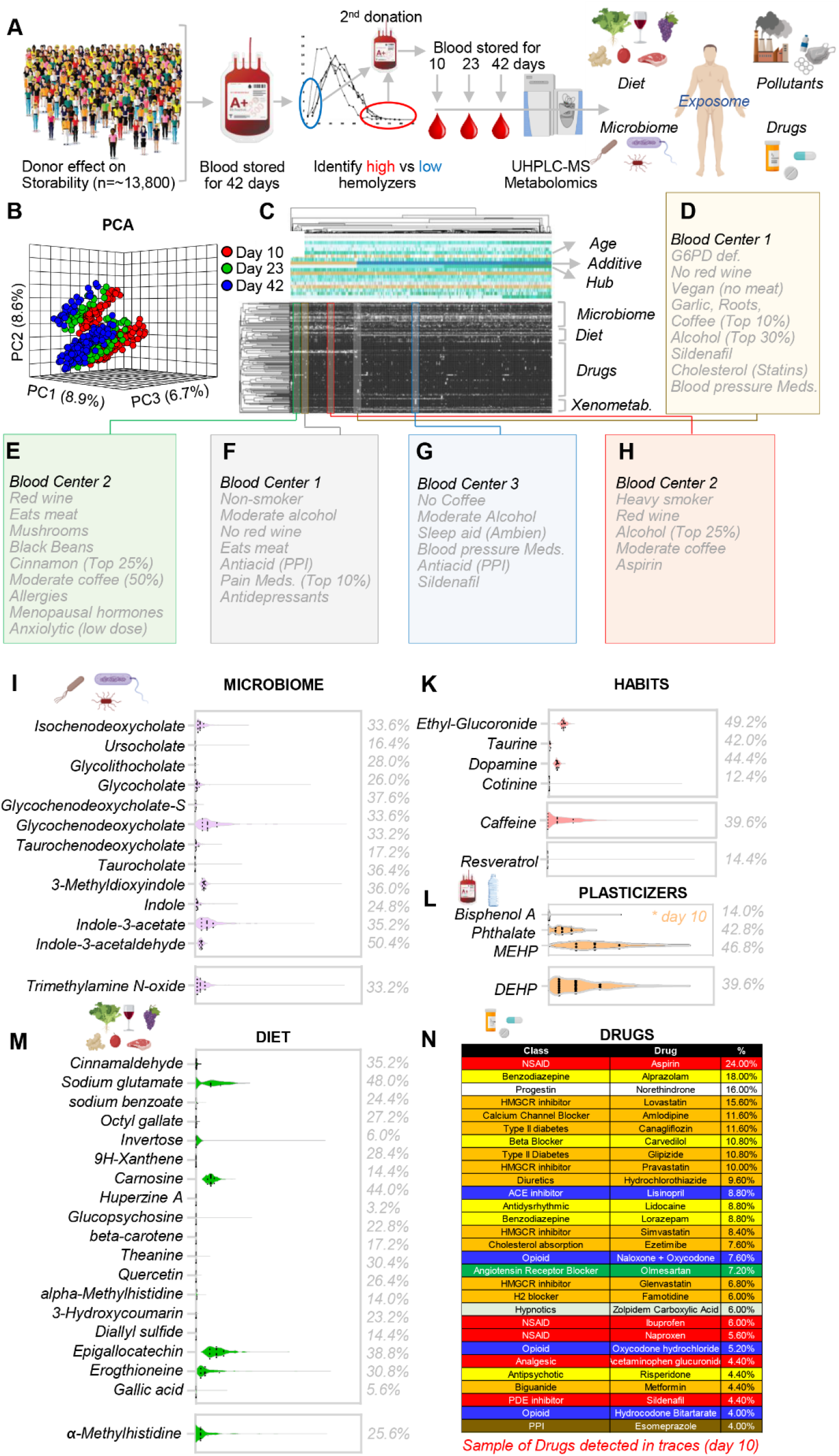
RBC exposome in the REDS-III RBC-Omics study. Within the framework of the Recipient Epidemiology and Donor Evaluation Study – REDS-III RBC-Omics study, 13,800 healthy donor volunteers were enrolled to donate a unit of whole blood that was processed into leukoreduced erythrocyte concentrates. Units were stored until the end of their shelf-life (42 days), when they were tested for the RBC propensity to hemolyze, spontaneously or following osmotic and oxidative stress. Donors in the 5^th^ and 95^th^ percentiles for hemolysis measurements were asked to donate a second unit of blood, which was sampled at storage days 10, 23 and 42 for metabolomics analyses of the RBC exposome, i.e. the compendium of small molecule metabolites derived from diet, the microbiome or other xenometabolites, such as pollutants or drugs (**A**). Unsupervised principal component analyses (PCA) of the exposome data clustered the subjects (dots) on the basis of storage duration (red, green, blue for days 10, 23 and 42, respectively (**B**). In **C**, hierarchical clustering analysis of blood donors (n = 250) at storage day 10 on the basis of exposome measurements (black to white: absent to high-abundance). The exposome informs on donor metabolic status and various habits, as highlighted in a few cases as highlighted from the HCA (**D-H**). In **I**-**N**, an overview of the most abundant exposome metabolites of microbial origin, habits (drinking alcohol or caffeinated beverages; nicotine exposure), plasticizers, dietary metabolites or common drugs (with % of day 10 stored blood donors’ samples in which traces of those drugs were detected).

**Figure 2.**
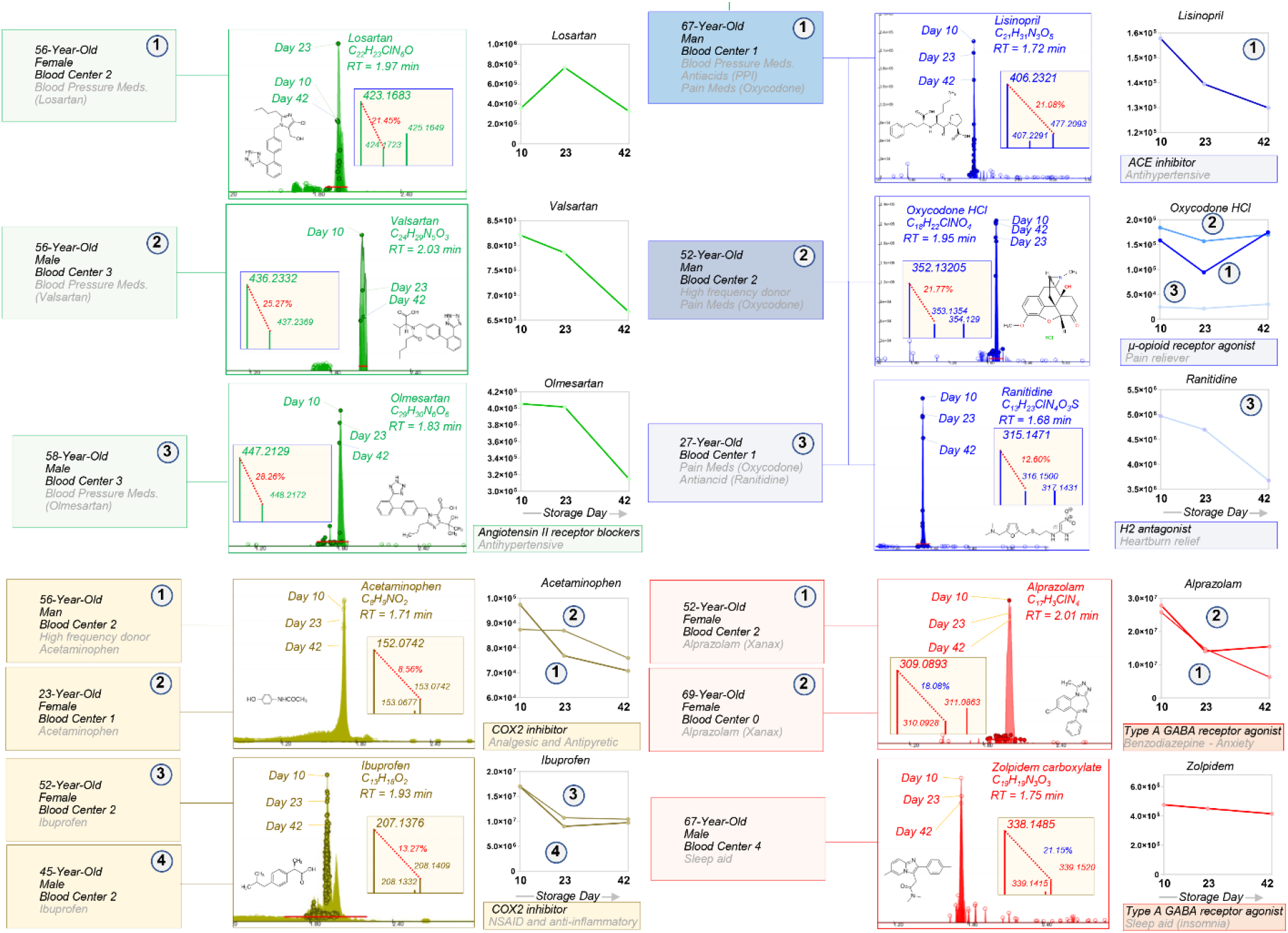
Exposome: traces of drugs were detected in a small subset of RBCs from healthy donor volunteers, including sartans (green), NSAID and other anti-inflammatory drugs (gold), antihypertensive drugs, pain medications (blue), antidepressants and sleep aids (red). Case studies are described in which the drugs were detected. For each metabolite we provide the original Extract Ion Chromatograms on the background of the whole population (599 samples), molecular formulae, retention times, mass spectra,^13^C abundance as percentage of the parent m/z peak, and time course levels of these metabolites.

Among all the units that tested positive for any drugs, the most significant deviation from the energy and redox metabolic phenotypes of the general population were noted in those subjects who tested positive for cholesterol medications (statins), blood pressure medications (angiotensin II receptor blockers) and antiacids (such as proton-pump inhibitors or antihistamine drugs) (**Supplementary Figure 3**).

### High-throughput metabolomics confirms a direct impact of FDA-approved drugs on RBC metabolism

In order to systematically validate the putative identifications and understand the potential impact of FDA-approved drugs on donor units, we performed a high-throughput metabolomics screening of human RBCs incubated with a panel of 1,366 FDA-approved drugs for 24h at 37ºC (**Figure 3.A-B**). To determine the metabolic impact of (classes of) drugs on RBCs, we calculated TSNE plots for 1,505 metabolomics samples of erythrocytes exposed to small molecule compounds (1,366 samples), vehicle (121 samples) or left untreated (18 samples – **Figure 3.C**). Upon normalization (**Supplementary Figure 4**), unsupervised analyses showed that vehicle and untreated samples clustered together. While we did not observe a clear clustering of small molecule compounds into specific classes, samples generally clustered by the magnitude of the drug perturbation (**Figure 3.C**). Indeed, small molecule drugs were found to alter the levels of erythrocyte metabolites that have been implicated in several pathologies and storage related changes^16^, including reduced glutathione, lactic acid, S-Adenosyl-L-methionine, and hypoxanthine (**Supplementary Figure 5**). In addition, machine learning models built using metabolites as predictors were able to accurately predict drugs for several drug classes/targets (i.e. bisphosphonates, anticholinergics, calcium channel blockers, adrenergics, proton-pump inhibitors, antimetabolites, selective serotonin reuptake inhibitors, and MTOR) suggesting that these drugs have a direct, conserved, and significant impact on RBC metabolism **(Supplementary Table 1**). To further understand impact on RBC metabolism, biclustering was used to identify “modules” (**Supplementary Table 1**), which are sets of drugs that induce similar changes to a similar set of metabolites. A total of 25 significant modules were identified due to perturbations by 331 small molecules affecting 151 metabolites (**Figure 3.D – Supplementary Table 1**). Of note, we also observed that RBC can metabolize a subset of the 1,366 drugs they were incubated with in the high-throughput screening, including primaquine to primaquine N-acetate and 5-hydroxy-desemthyl-primaquine (active metabolite – **Supplementary Figure 6.A**); tenofovir disoproxil to tenofovir metabolites (**Supplementary Figure 6.B**) and doxorubicin to doxorubicinol (**Supplementary Figure 6.C**).

**Figure 3.**
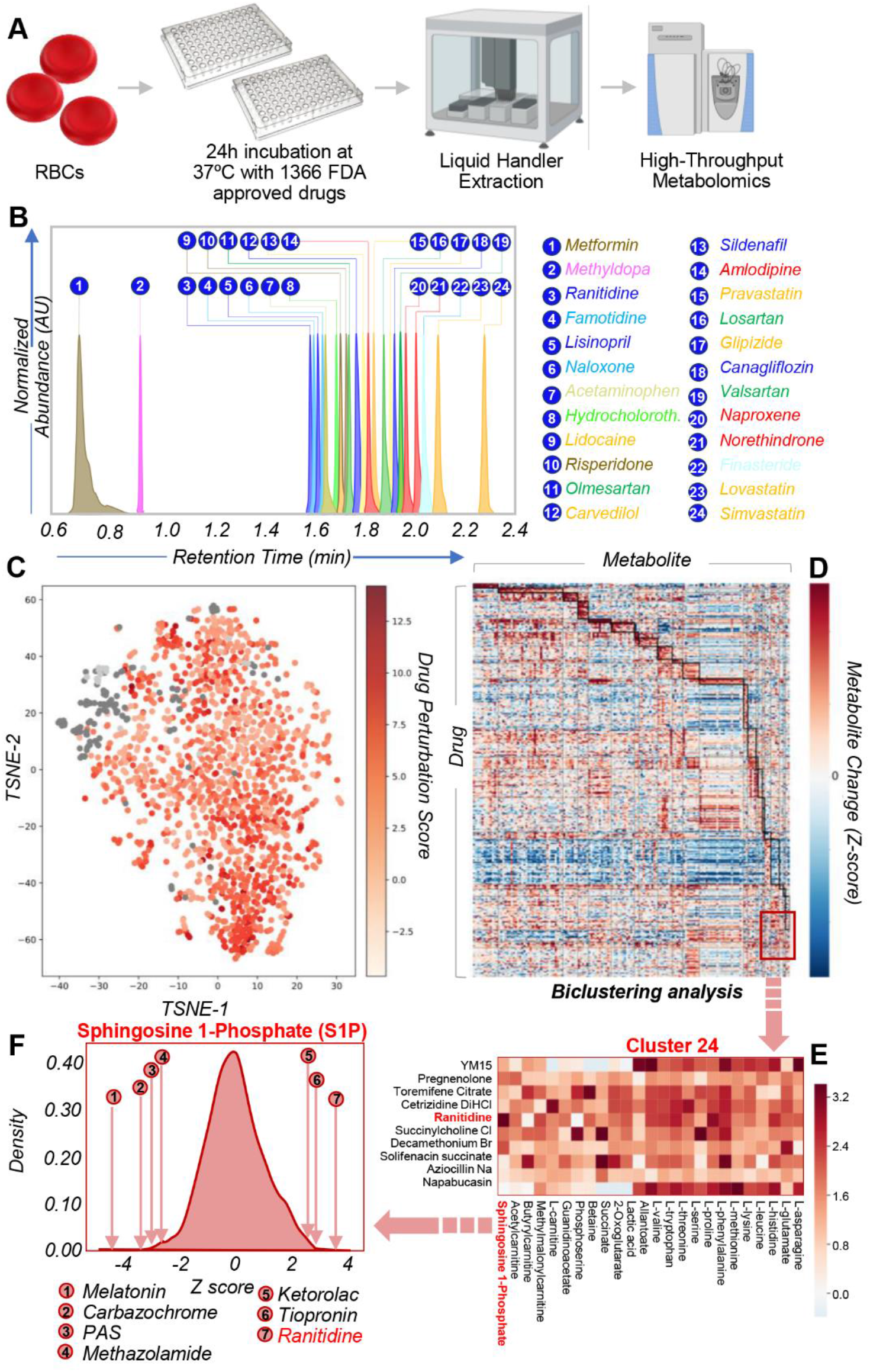
High-throughput metabolomics screening of human RBCs incubated with 1366 FDA-approved drugs for 24h at 37ºC (A). In **B**, retention times for most commonly identified drugs in the REDS III cohort were validated in the high-throughput screening (y-scale are Arbitrary unit – normalized across all drugs). In **C**, TSNE plot of 1507 metabolomics samples of erythrocytes exposed to small molecule compounds (1368 samples), vehicle (121 samples, dark grey), or left untreated (18 samples, light grey). Vehicle and untreated samples cluster together. Though there is not a clear clustering of small molecule compounds into specific groups, they generally cluster by drug perturbation Z-score. In **D**, Heat map of subset of the metabolomics data of erythrocytes exposed to small molecules. Biclustering was used to identify “modules” which are sets of drugs and set of metabolites that have similar changes. Modules are shown in dark black boxes. 25 total modules were identified due to perturbations by 331 small molecules affecting 151 metabolites. Identified modules and associated small molecules and metabolites are provided in Data SX. In **E**, a highlight of metabolites and drug in cluster 24 from the biclustering analysis. In **F**, effects of small molecule drugs from the high-throughput screening on the metabolite sphingosine 1-phosphate (S1P – one of the metabolites most significantly affected in cluster 24). Four compounds that decrease S1P the most and three drugs that increase S1P the most are shown. Melatonin is known to have an effect on the S1P signaling pathway. Carbazochrome is a hemostatic agent. 4-aminosalicylic acid (PAS) is an antibiotic for tuberculosis but is contraindicated in individuals with G6PDH deficiency as it induced hemolysis. Methazolamide is a carbonic anhydrase inhibitor. Ranitidine is an antihistamine. Histamine is known to affect S1P signaling.

### Ranitidine promotes increases in RBC Sphingosine 1-phosphate

To validate some of the findings from the high-throughput screening, we focused on cluster 24 from the biclustering analysis (**Figure 3.E**). This cluster included drugs with a significant effect on RBC markers of storage quality,^41^ e.g. lactic acid (marker of glycolysis – the main energy pathway in RBCs), acyl-carnitines (markers of membrane lipid homeostasis^42^), amino acids (marker of ion and protein homeostasis^10^), carboxylic acids (2-oxoglutarate and succinate – markers of altered homeostasis of reducing equivalents^43^) and sphingosine 1-phosphate (S1P), a major regulator of RBC glycolysis and function (i.e., oxygen off-loading) under physiological and pathological conditions.^35,44,45^ Of note, a subset of drugs were found to significantly decrease (e.g., melatonin, carbazochrome, 4-aminosalicylic acid – PAS, methazolamide) or increase (ranitidine, tiopronin and ketorolac) RBC S1P levels by over 4 standard deviations from the mean (Z-score normalized values) (**Figure 3.F**). Interestingly, melatonin has been reported to inhibit sphingosine kinase 1 (Sphk1)^46^ – the rate-limiting enzyme of S1P synthesis in RBCs. Ranitidine is an antihistamine (histamine H2 receptor antagonist), which is relevant in that histamine has been reported to positively regulate Sphk1 activity.^47^

### Detection of Ranitidine is associated with elevated S1P metabolism and S1P-regulated glycolysis in stored RBCs from the REDS III RBC Omics study

Only one subject tested positive for ranitidine in the recalled REDS III RBC Omics donor cohort(**Figure 4.A**), but samples from this donor ranked in the top 13% S1P levels of all the 599 samples tested (**Figure 4. B-C**). We have previously shown that RBC S1P promotes glycolysis by stabilizing the tense deoxygenated state of hemoglobin, which in turn outcompetes glycolytic enzymes that are otherwise bound to and inhibited by the N-terminus of band 3 (model summarized in **Figure 4.D**).^35,44,45^ Consistently, RBCs from the ranitidine positive unit were characterzied by end of storage lactate levels in this subject that ranked 2^nd^ out of all 599 samples tested in this study(**Figure 4.E**). Given the structural similarity between ranitidine and S-Adenosyl-Homocysteine (SAH – **Figure 4.F**), we hypothesized and observed an association between the detection of ranitidine and the levels of SAH and related metabolites (SAM and SAM/SAH ratios – **Figure 4.F**), metabolites critical in oxidant stress-induced isoaspartyl protein damage-repair in the stored erythrocyte.^5^ Consistent with our prediction based on the high-throughput screening, RBCs from the subject on ranitidine were characterized by decreases in the levels of several metabolic markers of the storage lesion (**Figure 4.G**), including markers of poor post-transfusion recovery hypoxanthine, arachidonic acid and 12-HETE (lowest levels across all 250 subjects enrolled in this study – **Figure 4.H**).^36,41^

**Figure 4.**
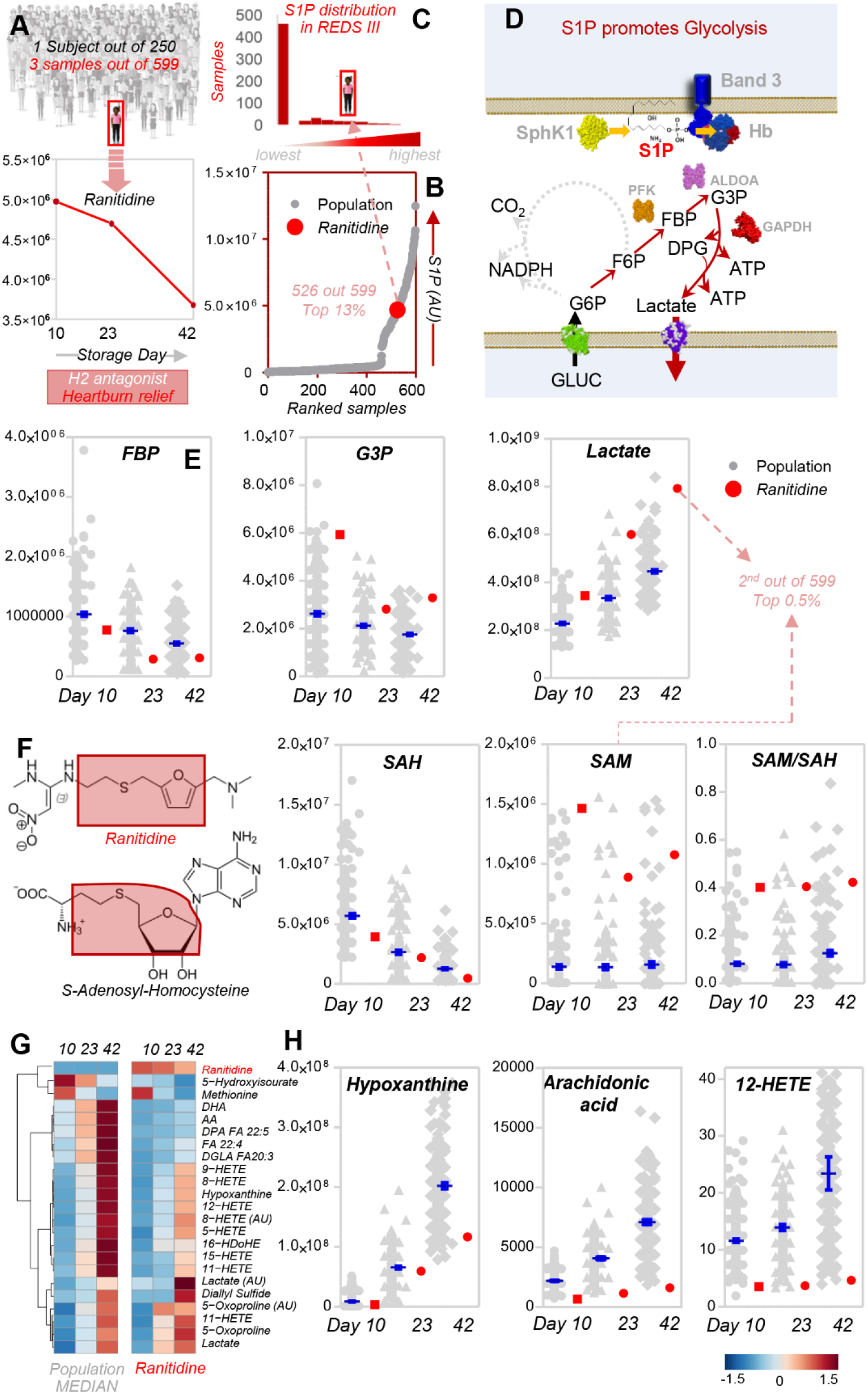
Ranitidine impacts S1P metabolism and S1P-regulated glycolysis in stored RBCs from the REDS III RBC Omics study. Only one subject was identified to be positive for ranitidine out of 599 subjects in the recalled REDS III RBC Omics donor cohort (**A**). S1P levels in RBCs from this subject at storage day 10, 23 and 42 ranked in the top 13% across all the 599 samples tested in this study (**B-C**). In **D**, a schematic representation of the mechanisms through which S1P levels in RBCs modulate glycolysis, as previously described in Sun et al. Nat Comm 2016. Consistently, faster consumption of early glycolytic intermediates (fructose bisphosphate – FBP) and increased levels of glyceraldehyde 3-phosphate (G3P) and the final byproduct of glycolysis, lactate were detected in the subject positive for ranitidine (**E**), with end of storage lactate levels rankind 2^nd^ of all the 599 samples tested in this study (Top 0.5%). Given the structural similarity between ranitidine and S-adenosylhomocysteine (SAH – **F**), we hypothesized and confirmed an interaction of this metabolite with the levels of SAH and related metabolites (SAM and SAM/SAH ratios – **F**). In **G**, heat map of the top 25 metabolites that change significantly between the subject positive for ranitidine at any storage day (10, 23 and 42) and the median of the rest of the population. The map highlighted that several metabolic markers of the RBC storag lesion are decreased in the stored RBCs from this subject, including markers of post-transfusion recovery hypoxanthine, arachidonic acid and 12-HETE (**H**).

### Ranitidine boosts S1P levels and glycolysis in a dose-response fashion in human and WT mouse RBCs, but not in Sphk1 KO mice

To test whether ranitidine was causally associated and not just correlated with improved energy and redox metabolism in RBCs, human RBCs (n = 3) were incubated with increasing doses of ranitidine in presence 1,2,3-^13^C_3_-glucose for up to 60 min (**Figure 5.A)**. At increasing doses of ranitidine (**Figure 5.B)** corresponded increasing levels of^13^C_3_-labeled 1,6-frucotse diphosphate (rate limiting step of glycolysis, plateauing at 100uM ranitidine – **Figure 5.C**) and^13^C_3_-lactate (**Figure 5.D**)– suggestive of increased fluxes through glycolysis. RBCs were also obtained from three WT and Sphk1 KO mice,^35^ prior to incubation with 100 uM ranitidine for 60 min at 37ºC (**Figure 5.E**). After confirming significant decreases in S1P levels in the Sphk1 KO mouse RBCs (**Figure 5.F**), we observed that ex vivo incubation with ranitidine promoted increases in the levels of several glycolytic intermediates, including glucose 6-phosphate (G6P), glyceraldehyde 3-phosphate, 2,3-Diphosphoglycerate (2,3-DPG), phosphoglycerate, and pyruvate. However, increases in the total levels of lactate were observed only in WT but not in Sphk1 KO mice following incubation with ranitidine. On the other hand, Sphk1 KO mice showed increased steady-state levels of ribose phosphate, suggestive of increased fluxes through the pentose phosphate pathway and altered redox homeostasis (e.g., glutathione, methionine) – consistent with the model in **Figure 4.D**. Finally, the total adenylate pools were increased in WT cells following incubation with ranitidine, but decreased in Sphk1 KO mouse RBCs (**Figure 5.F**). Since different mouse strains respond differently to the energy and redox storage lesion,^36^ storage of RBCs from mice with poor (FVB n = 3) or good (B6 n = 3) storage quality in presence of ranitidine (0, 50, 100 or 200 uM) resulted in significant improvements in RBC storage quality, especially at 100 uM concentration (**Figure 5.G**) and increases in RBC S1P levels in both strains (**Supplementary Figure 6**).

**Figure 5.**
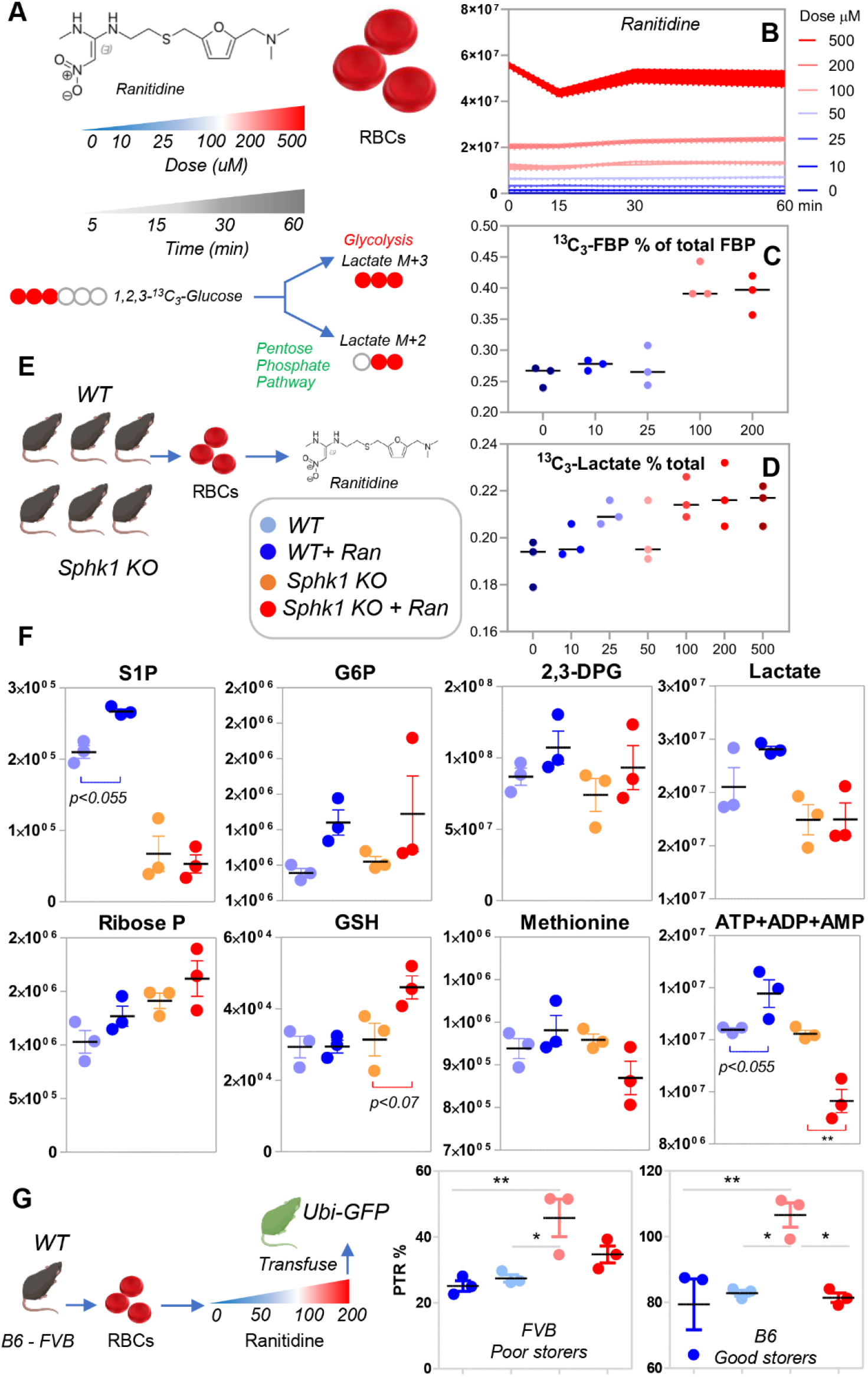
Ranitidine boosts glycolysis in a dose-response fashion in human RBCs. Human RBCs (n = 3) were incubated with increasing doses of ranitidine in presence 1,2,3-^13^C_3_-glucose for up to 60 min, as detailed in **A**. At increasing doses of ranitidine (**B**) corresponded increasing levels of^13^C_3_-labeled 1,6-frucotse diphosphate (rate limiting step of glycolysis, plateauing at 100uM ranitidine – **C**) and^13^C_3_-lactate (**D)**– suggestive of increased fluxes through glycolysis but not the Rapoport Luebering shunt. RBCs were also obtained from three WT and Sphk1 KO mice, prior to incubation with 100 uM ranitidine for 60 min (**E**). After confirming significant decreases in S1P levels in the Sphk1 KO mouse RBCs (**F**), we observed that ex vivo incubation with ranitidine promoted increases in the levels of several glycolytic intermediates, including glucose 6-phosphate (G6P), glyceraldehyde 3-phosphate, 2,3-Diphosphoglycerate (2,3-DPG), phosphoglycerate, and pyruvate. However, increases in the total levels of lactate were observed only in WT but not in Sphk1 KO mice following incubation with ranitidine. On the other hand, Sphk1 KO mice showed increased steady-state levels of ribose phosphate, suggestive of increased fluxes through the pentose phosphate pathway and increased NADPH-dependent recycling of glutathione. This observation is consistent with the observed increases in the levels of reduced glutathione in Sphk1 KO mice after incubation with ranitidine. On the other hand, these mice were characterized by lower levels of another antioxidant, methionine. Finally, the total adenylate pool (high energy phosphate compounds including adenosine tri-, di- and mono- phosphate – ATP+ADP+AMP) were increased in WT cells following incubation with ranitidine, but decreased in Sphk1 KO mouse RBCs (**F**). Color-codes for panels **E** and **F** are explained by the legend in the center of panel **E**. Storage of RBCs from mice with poor (FVB n = 3) or good (B6 n = 3) storage quality in presence of ranitdine (0, 50, 100 or 200 uM) resulted in significant improvements in RBC storage quality at 100 uM concentration (**G**). Color codes in panel **G** are consistent with ranitidine doses as per the stylized color legend at the center of this panel (explicitly described in panel **B**).

**Figure 6.**
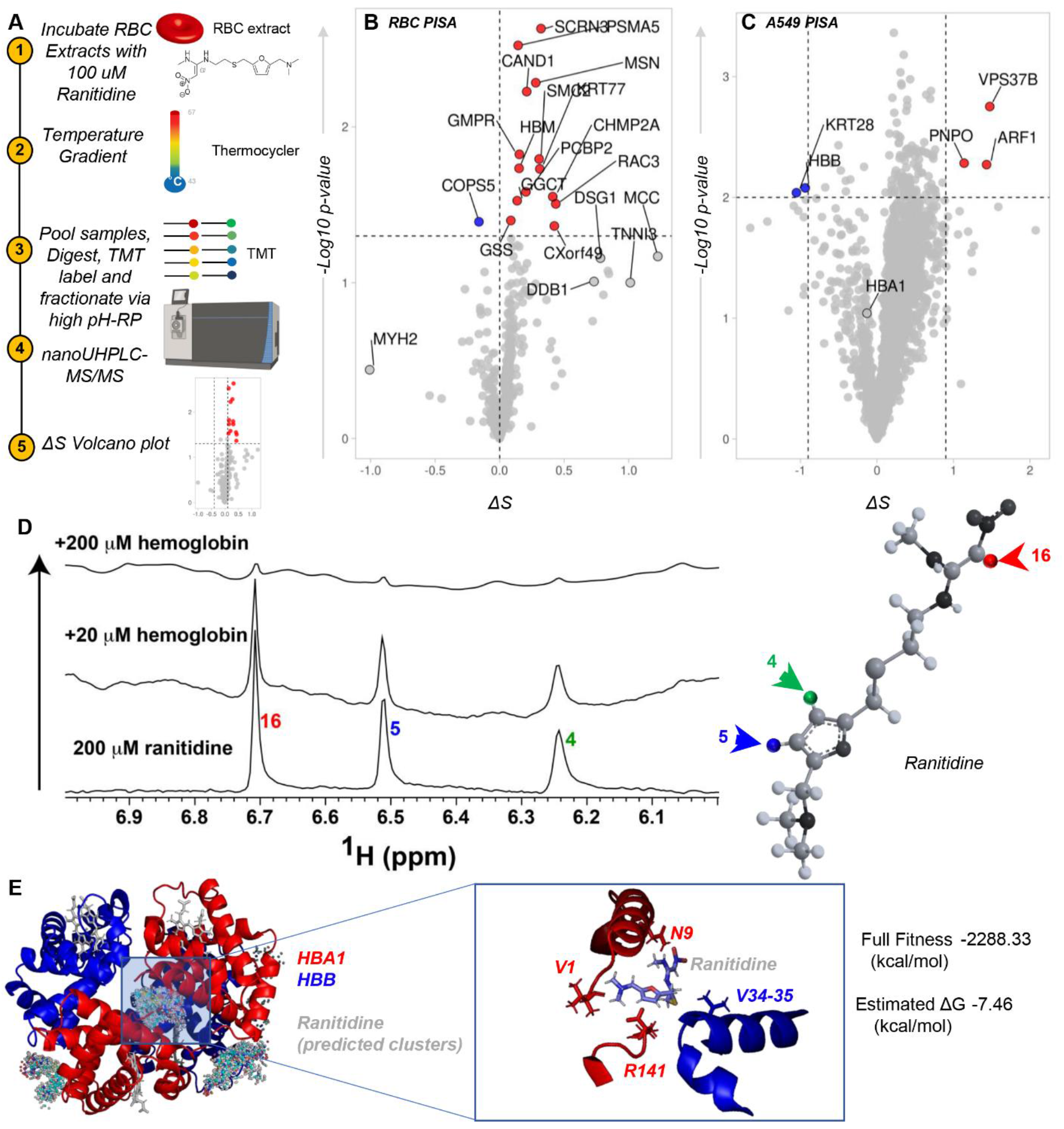
Proteome Integral Solubility Alteration (PISA) assay in RBCs and A549 epithelial cells in presence of 100 uM ranitidine. In **A**, an overview of the experimental design. In **B** and **C**, volcano plots show the proteins that are significantly stabilized (red) or destabilized (blue) by ranitidine in RBC and A549 extracts, respectively. In **D**, a 1D NMR 1H spectrum was collected of 200 uM ranitidine alone and in the presence of 20 uM hemoglobin and 200 uM hemoglobin at 25ºC. 48 scans were collected on a Varian 900 using the BioPack water sequence implemented with wet water suppression. All spectra are shown at the same vertical scale. The disappearance of peaks means that the resonances of ranitidine – the smaller molecule – are relaxing faster due to binding to the larger molecule – hemoglobin. On the right hand side of the panel, an overview of the ranitidine structure with highlighted in red, blue, green the proton responsible for the resonances highlighted in the spectra to the left. In **E**, docking of ranitidine with the deoxyhemoglobin tetramer (1a3n.pdb) was calculated in silico with SwissDock. All possible dockings are shown in the left hand portion of the panel. On the right hand side of the panel, zoom in into the conformation with the lowest estimated Gibbs free energy and best fitness, at the interface between the two alpha globin chains (at the N-term and C-term of either HBA1 chain – red).

### Proteome Integral Solubility Alteration assay suggests a potential interaction of ranitidine with hemoglobin beta

Finally, to provide a preliminary understanding of the potential mechanism underlying our observations, we performed PISA assays to determine ranitidine targets in RBCs (experimental design and volcano plot in **Figure 6.A-B**, respectively). Interestingly, results identified a small number of potential ranitidine interactors, including hemoglobin subunits (e.g., hemoglobin subunit mu – HBM) and several enzymes involved in redox homeostasis (e.g., Gammaglutamylcyclotransferase – GGCT; glutathione synthetase – GSS), GMP metabolism (GMP reductase – GMPR; Ras-related C3 botulinum toxin substrate 3 – RAC3), structural homeostasis (e.g., moesin – MSN), vesiculatin (secernin 3 – SCRN3) and protein degradation (Cullin-associated NEDD8-dissociated protein 1 – CAND1; proteasomal subunit A5 – PSMA5) (**Figure 6.B**). However, acknowledging the technical limitations of these experiments in RBCs, owing to the overwhelming abundance of hemoglobins, we decided to repeat the experiment in A549, an epithelial alveolar cell line that has been reported to express hemoglobins^48^. As a result (**Figure 6.C**), a cleaner readout was obtained, highlighting a destabilizing effect of ranitidine on hemoglobin beta (HBB) chain. Finally, 1D NMR 1H studies were performed to test the predicted interaction of ranitidine with hemoglobin (**Figure 6.D**). Results indicate a clear disappearance of peaks assigned to ranitidine protons, suggesting that the resonances of ranitidine, the smaller molecule, are relaxing faster due to binding to the larger molecule, hemoglobin tetramer – in a dose-dependent fashion (0, 20 and 200 uM hemoglobin). We thus used the software SwissDock^49^ to perform *in silico* docking prediction of the interaction between hemoglobin (pdb: 1a3n) and ranitidine. Deoxygenated hemoglobin was selected rather than the oxygenated form because of the impact of ranitidine on S1P observed in this study and prior evidence of S1P interaction with deoxyhemoglobin.^44^ Results from this computation are shown in **Figure 6.D** – with all the possible docking conformations mapped in the left hand panel. Zooming in into the fitting conformation with the lowest predicted Gibbs free energy (right hand panel – **Figure 6.D**), ranitidine was predicted to sit at the interface between the N-term (V1 and N9) and C-term (R141) of the two alpha hemoglobin chains, respectively, and in proximity to valines 34 and 35 of HBB.

## Discussion

In the present study, we provide the first comprehensive description of the blood donor exposome – i.e., the compendium of small molecule metabolites that derive from exogenous exposures, including but not limited to diet, xenometabolites (e.g., drugs), habits (e.g., smoking, drinking, etc.) or blood processing. Previous studies focused on the metabolic impact of specific exposures, such as donor habits (e.g., smoking,^24,25^ consumption of alcohol,^50^ coffee^51^ or taurine^52^), or blood processing (e.g., plasticizers, storage additives, leukofiltration). As such, here we mostly focused on an as of yet unexplored aspect of RBC storage biology: the presence and potential impact of drugs on stored RBC metabolism.

Since blood donor deferral is only limited to a shortlist of drugs impacting coagulation cascades (e.g., blood thinners) and antibiotics (full list available at this link)^53^, it is expected – but to the best of our knowledge unreported – that routine blood donors would be exposed to off-the-counter or prescription drugs at a rate comparable to the general population. Personalized medicine efforts worldwide are increasingly challenging the very concept of “healthy” control subjects in medicine. Here we showed that this caveat holds true in a population of healthy donor volunteers, a demographic group that has progressively aged over the past two decades and is thus statistically more likely to consume drugs that impact RBC metabolism and storability (e.g., blood pressure medications, statins, biguanides, aspirin, antiacids). From the present analysis it is not possible to determine (i) if those drugs were present only in traces or in concentrations of potential relevance to the recipient; (ii) how close to the blood donation the drugs had been taken; (iii) whether the drugs are enriched in RBCs or in the residual plasma in the supernatants, which accounts for ∼10% of the storage medium in which the RBCs are resuspended. Interestingly, some of the drugs like sildenafil or progestin (birth control) were detected in donors with a demographic consistent with their clinical target (e.g., males over 65 or reproductive age females, respectively), providing internal validation for the present analysis. These observations were accompanied by the detection of some metabolites of these drugs in the blood units, consistent with the observed decrease in the levels of most detected drugs as a function of storage duration in longitudinal samples. While drug metabolites may have ended up in the unit upon metabolism from other organs prior to donation (e.g., liver), here we provide preliminary evidence that incubation of leukocyte-filtered RBCs with a drug library of 1,366 FDA approved drugs afforded the detection of some drug metabolites, suggestive of an active drug metabolism for a subset of small molecule chemicals in the mature erythrocyte. This observation expands on recent reports on the metabolism of antiretrovirals (e.g., tenofovir)^15^ and antimalarials^14^ in the mature erythrocyte, a phenomenon perhaps explained by the activity of RBC abundant cytochrome b5 reductase or other diaphorases in this cell.^10^

While future studies will further delve into the role of RBC in drug metabolism, here we leveraged a high-throughput drug screening to confirm that ∼65.1% of FDA-approved drug in the library we tested had a significant effect on RBC metabolism. This is relevant in that it supports the hypothesis that drug consumption may represent an unappreciated contributor to the metabolic heterogeneity of stored blood units. Our observation does not necessarily hold a negative connotation, in that some drugs have been observed to favor RBC energy metabolism and decrease oxidant stress markers, including markers of poor storage quality and post-transfusion performances.^36,41,54^ Indeed, we provide evidence of a metabolic impact of ranitidine, a histamine H2 receptor antagonist that is prescribed and off-the-counter drug in the treatment of gastrointestinal diseases related to gastric acid hypersecretion. We show that detection of ranitidine is associated with increases in energy metabolism and decreased oxidant stress in the REDS III-RBC Omics recalled donor cohort. These observations were validated in human RBCs in vitro and mouse RBCs ex vivo, in which ranitidine promoted increases in the levels of S1P and fluxes through glycolysis in WT mice, but not in mice lacking the expression of Spk1 – the rate-limiting enzyme of S1P synthesis. Mouse RBCs stored in presence of ranitidine were characterized by higher S1P levels – which normally decrease as function of storage under blood bank conditions – and increased post-transfusion recoveries, i.e., the percentage of transfused RBCs that still circulate 24h after transfusion – in two different mouse strains. In prior work, we have shown that RBCs up-regulate S1P synthesis via Sphk1 to counteract hypoxia at high altitude or in sickle cell disease.^35,44^ By stabilizing the tense deoxygenated state of hemoglobin, S1P promotes the displacement and activation of glycolytic enzymes from the N-terminus of band 3, which in turn favors metabolic fluxes through glycolysis.^35,44^ In analogy to this model, here we provide evidence (PISA, *in silico* docking and NMR) of a potential interaction between ranitidine and hemoglobin.

In conclusion, we provide evidence of high rates of detection of drugs, drug metabolites and other exposome compounds in stored RBC units. We also show an impact of drugs (65% of 1,366 tested) on RBC metabolism and offer an example of the translational clinical relevance of our findings in transfusion medicine. Through a combination of high-throughput metabolomics and machine learning elaborations, the data generated in this study helped to formulate a specific hypothesis that we further tested mechanistically (i.e., ranitidine improves RBC energy and redox metabolism, storage and post-transfusion performances), and several others that will fuel years of follow-up investigations.

## Data Availability

All the raw data are provided in the Supplementary Table of this manuscript.

## Acknowledgments

Research reported in this publication was funded by the NHLBI Recipient Epidemiology and Donor Evaluation Study-III (REDS-III), which was supported by NHLBI contracts NHLBI HHSN2682011–00001I, –00002I, –00003I, –00004I, –00005I, –00006I, –00007I, –00008I, and –00009I, as well as funds from the Institute of General and Medical Sciences (RM1GM131968 to ADA and KCH), the Boettcher Webb-Waring Investigator Award (ADA), and R01HL146442 (ADA), R01HL149714 (ADA), R01HL148151 (ADA, JCZ), R21HL150032 (ADA), and T32 HL007171 (TN) from the National Heart, Lung, and Blood Institute. The content is solely the responsibility of the authors and does not necessarily represent the official views of the National Institutes of Health. The authors would like to express their deep gratitude Dr. Simone Glynn of NHLBI for her support throughout this study, the RBC-Omics research staff at all participating blood centers and testing labs for their contribution to this project, and to all blood donors who agreed to participate in this study..

## Authors’ contributions

TN, DS, ADA performed metabolomics analyses; AB, BOP, ADA performed data analysis. LJD, SK, MPB performed blood donor studies. AH, AS, YX, JCZ performed animal studies. AI, KCH performed PISA studies. JSR, EZE performed NMR studies.

